# Non-Contact Molecular Chemical Imaging Assessment of Tissue Congestion: A Validation Study Compared to MRI

**DOI:** 10.1101/2022.11.01.22281818

**Authors:** Adam E Saltman, Richik N Ghosh, Peter M. Eckman, Michael Chappuis, Jeremy D Collins, Pamela Morley, Holly Mazis, Robert Schweitzer, Maurice Enriquez-Sarano, Shawna K Tazik, Heather Gomer, Matthew Nelson, Patrick J Treado

## Abstract

**Background:** Heart failure (HF) is a healthcare problem burdening patients, care teams, and health care systems. A relentless, downward spiral of decompensation, hospital admission, and functional deterioration is challenging to break, partly because of the lack of an objective, inexpensive tool to aid the clinician’s physical examination. We studied performance of a shortwave infrared (SWIR) molecular chemical imaging (MCI) tool to measure relative tissue congestion (TC) in HF patients’ shins, compared to MRI Dixon sequence TC measurements as ground truth.

**Methods:** Forty-seven (47) subjects underwent paired SWIR MCI and MRI measurements of their lower extremities. Thirty-six (36) subjects were hospitalized with decompensated HF while 11 healthy outpatients served as controls. A partial least squares (PLS) regression model was trained to ingest the SWIR MCI spectra and produce a CardioVerification Index (CVI) that mirrored MRI measurements.

**Results:** The SWIR MCI model reflected MRI TC measurements accurately for all subjects with a 0.743 linear correlation coefficient. Surprisingly, the MRI results identified a significant fraction of non-HF subjects with elevated TC levels, so a sub-analysis was performed on “Low TC” subjects with low and undetectable levels of extremity edema. This subpopulation’s linear correlation between MRI and CVI was 0.674. A logistic classifier model differentiated HF from non-HF subjects; the area under the receiver operating characteristic curves was 0.906 for the entire subject group and 0.821 for the Low TC subpopulation.

**Conclusion:** SWIR MCI techniques can be used in a TC-measuring tool that replicates MRI TC measurements. Clinicians could use such tools to help guide therapy for HF patients during in-hospital management of acute HF decompensation and for outpatient monitoring. SWIR MCI’s ability to identify elevated TC in otherwise normal subjects may also meet the unmet need for a novel, widely applicable HF screening tool.

## Introduction

Heart failure (HF) is a leading cause of morbidity and mortality, and a major burden for patients, healthcare providers, and healthcare systems. The prevalence of HF in the US is 6.2 million, and by 2030, expected to be 8 million.^1^ Annual estimates for HF care per patient range from $868 in South Korea, $24,383 in the US, to $25,532 in Germany, with a worldwide average lifetime cost of $126,819.^2-3^ Furthermore, HF patients live in an unrelenting cycle of deterioration, with variable stretches of stability punctuated by episodes of decompensation requiring emergency department visits often resulting in hospital admissions; each revolution of this cycle worsens patient morbidity and mortality.^4,5^

The American Heart Association and the American College of Cardiology have staged HF from A to D, based on clinical symptomatology.^6^ Stage A and B patients have no signs or symptoms of HF. Stage A patients have only risk factors for HF, and treatment is aimed at risk reduction. Stage B patients have structural heart disease, and treatment is directed appropriately (*e.g*., ACE inhibitors for those with a reduced ejection fraction). Patients in stages C and D present with signs of HF (*e.g*., shortness of breath, leg swelling); the difference between them is that stage C patients can be managed effectively with medications and certain implantable devices, while stage D patients require advanced, non-pharmacological treatments in addition to pharmacotherapy.

Regardless of HF stage, patient functional status changes along a common pathway of increased tissue congestion (TC). In the lungs, congestion is experienced as pulmonary edema and shortness of breath. In the periphery, congestion is experienced as peripheral edema and swelling. Although not completely understood, it appears that congestion occurs simultaneously in the two regions, likely two or three weeks before gross clinical decompensation and hospitalization.^7^ Therefore, key goals in approaching HF patients are to (1) find patients as early in the disease, preferably before they show clinical signs of TC, and (2) monitor them closely for increasing TC, so that prompt intervention can keep them out of the hospital.

Our ability to diagnose patients early (stage B) depends upon screening an asymptomatic population, which despite many attempts remains a vexing, unsolved problem.^8,9^ Our ability to diagnose and manage patients in stages C and D depends upon appraisal of the patient’s history, symptoms, and physical signs. Unfortunately, appreciation of such signs as peripheral edema and weight gain often come later in the disease, are imprecise, have high interobserver variability and poor sensitivity.^10^ Significant advances could come from use of a device that provides early TC detection before onset of HF symptoms (screening) or worsening of existing HF symptoms (monitoring). By detecting these physiological changes days to weeks before acute HF decompensation events, such a device would have a significant positive impact on managing HF patients.

For this study, we compared CardioVere® (CV), a non-invasive, non-contact, optical device that uses shortwave infrared (SWIR) molecular chemical imaging (MCI), to magnetic resonance imaging (MRI) and physical examination for detecting and tracking changes in TC. The study had four objectives:

- To determine whether CV can accurately differentiate non-HF subjects from acutely decompensated HF subjects
- To apply in a novel way the MRI Dixon sequence method of water measurement in quantifying TC in HF patients’ shins
- To assess CV’s performance in quantifying TC, compared to MRI as the ground truth
- To quantify the utility of the physical examination, represented by the grading of pitting edema, as a method of TC measurement

## Materials and Methods

### Subject selection

Study subjects were recruited from inpatients at Abbott Northwestern Hospital who had acutely decompensated HF (ADHF). Control (non-HF) subjects were recruited from the Minneapolis Heart Institute clinical coordinator staff and the manuscript authors (M.C. and M.S.), none of whom carried any diagnosis, signs, or symptoms of HF.

All subjects underwent single 6-point Dixon LiverLab sequence MRI imaging and CV imaging on the lower extremities.^10–17^ Each subject was imaged once, with MRI and CV imaging events taking place within 4 hours of each other. The ADHF subjects were imaged at a random time during their hospitalization, and the control subjects were imaged during an unrelated office visit. Informed consent was obtained from all study participants. Enrolled subjects were allocated a deidentified alphanumeric study identifier under which all imaging and metadata were collected. The study was approved by the Institutional Review Board of Advarra and Allina Abbott Northwestern Hospital.

#### Inclusion criteria

- Age ≥ 18 years
- (Study group) Acutely decompensated heart failure, any ACC/AHA stage or NYHA class
- (Control group) No known diagnosis of HF
- Able and willing (or health care proxy able and willing) to give consent

#### Exclusion criteria

- Skin lesions that would affect CV use
- Inability to tolerate MRI examination
- HF requiring emergency treatment
- Unstable hemodynamics
- Inability to remain recumbent for CV and/or MRI examination
- Active exacerbation of deep vein thrombosis
- Pregnancy

### Physical examination evaluation

Variability in the physical examination was determined by calculating inter-rater reliability for pitting edema (PE) grading. Two experienced clinicians independently assessed PE in the ADHF subjects at the same level on the shin and graded its presence and severity from 0 to 4+ using standardized definitions.^18^ The clinicians performed their grading within 20 minutes of each other, except for one subject, where 2 hours elapsed. Each clinician was blinded to the other’s determination. Interobserver reliability was expressed as Cohen’s kappa.^19^

### Water fraction measurement using the MRI Dixon sequence method

The Dixon method is an MRI sequence based on the differences between fat and water protons’ resonance frequencies that decomposes the signal into two separate water-only and fat-only image maps enabling direct image-based water and fat quantitation.^11,12,15^ Dixon sequencing does not require injection of imaging agents to visualize fat and water distributions. The presence of water in the Dixon sequence generated water-only images are clearly seen (Figure 1A).

**Figure 1:** Detection and measurement of interstitial water in shins by the MRI Dixon method

**Figure 1A:**
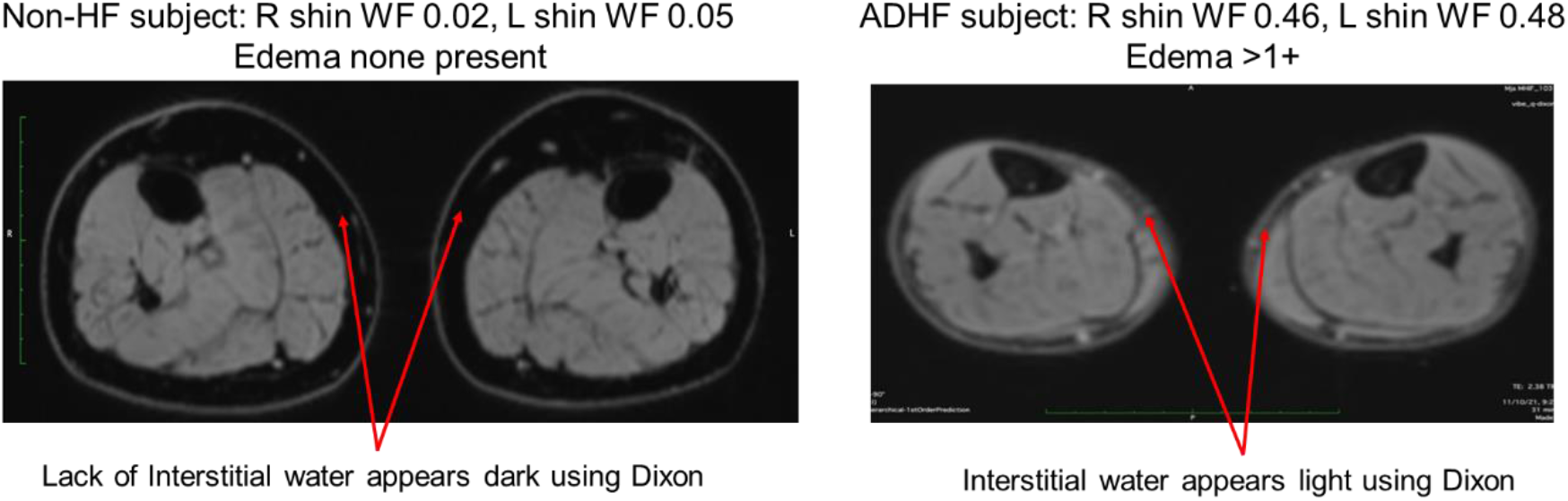
Detection of interstitial water in shins by MRI Dixon method in a non-HF subject (left) and an HF subject (right). In the water-only axial views of the shin periphery that are shown, the lack of water appears dark whereas the presence of water appears lighter.

For this study, using a Siemens MRI system with a 1.5T magnet, an MRI scan of the lower extremity was performed on each subject using the 6-point Dixon LiverLab sequence (Siemens USA, Cary, NC).^10^ On average, this yielded approximately thirty-six (36) 4mm thick axial images per subject. Two slices were utilized per shin, one proximal to the knee and the other proximal to the ankle. Using LiverLab MRI software, two regions of interest (ROI) in the interstitial space beneath the skin were defined per slice (Figure 1B). Due to variability in the lateral readings, only the medial ROIs were used for quantitation. Overall, four MRI ROIs were analyzed per subject (2 shins × 2 slices per shin).

**Figure 1B:**
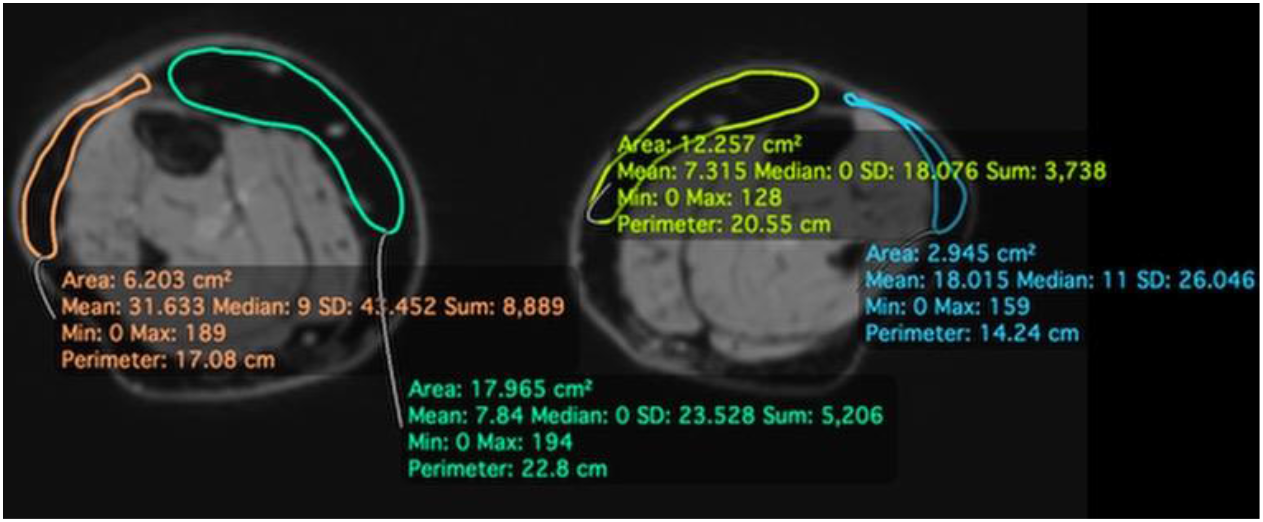
Measurement of interstitial water in periphery of shins by MRI Dixon method. A region of interest (ROI) is drawn in the peripheral region below the skin (water-only image shown), and the signal from the water-only and fat-only images in the ROI are measured to give the water fraction.

From the water-only and fat-only images, the water fraction (WF) was defined as the relative, quantifiable amount of excess water contained within the interstitial tissue. WF is determined from the signal in the water-only image ROI divided by the sum of the water-only and fat-only signals in the same ROI:

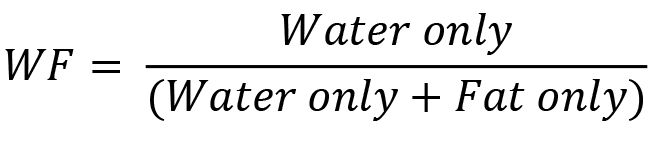

### Water fraction measurement using CardioVere

CardioVere (CV) is a non-invasive, non-contact, optical device that uses SWIR MCI to measure the water content of living tissues.^20^ Figure 2A shows the imaging system and major components, and Figure 2B, the SWIR MCI workflow.^20^ Briefly, MCI integrates digital imaging with spectroscopy to provide both spatial and molecular information about a sample. The technology illuminates the subject’s shin with broad-band white light and acquires a hypercube which contains a collection of high-definition, reflected light, wavelength-resolved images in 5 nm increments over the SWIR spectral range (wavelengths: 900 – 1700 nm). The sensor head unit (SHU) collects the hyperspectral images via an articulating arm, enabling its positioning normal to each limb at ∼40 cm working distance. Illumination of the 318 mm × 254 mm field of view is by a light housing equipped with halogen lighting attached to the SHU. Once positioned, the operator controls data collection via the Spectral Kitchen® software (ChemImage Corp., Pittsburgh PA) which runs on the device’s computer.

**Figure 2:** SWIR MCI Workflow

**Figure 2A.**
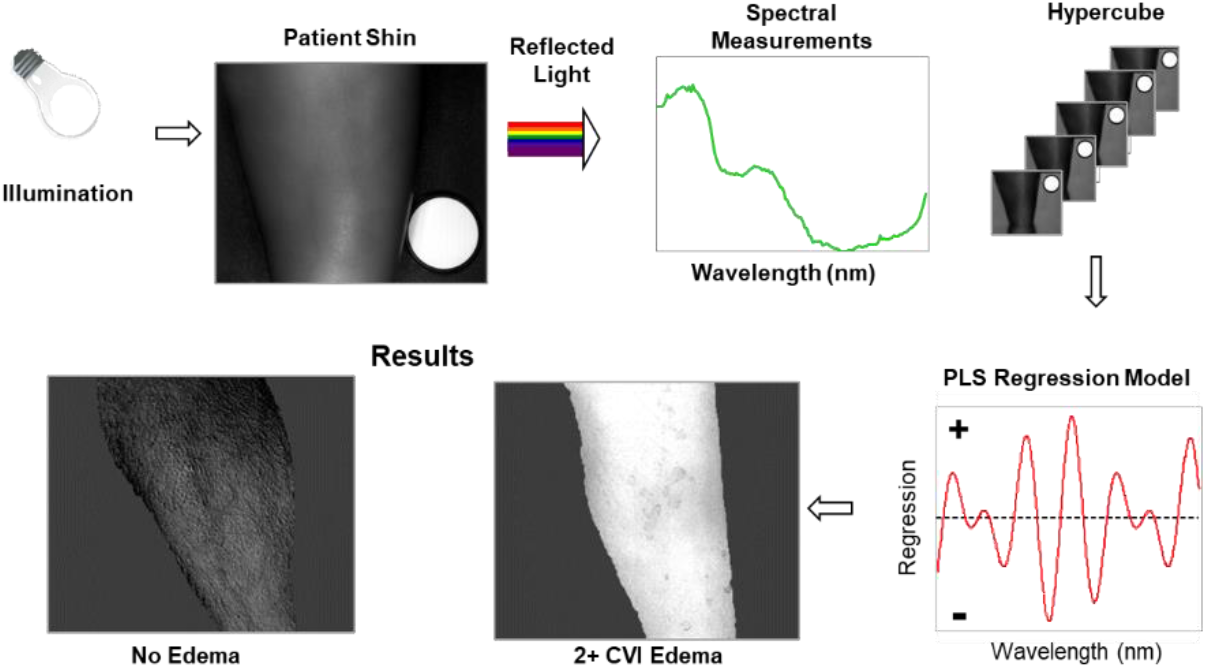
Left: SWIR MCI device with main components. Right: SWIR MCI imaging of a healthy subject’s shin.

**Figure 2B:**
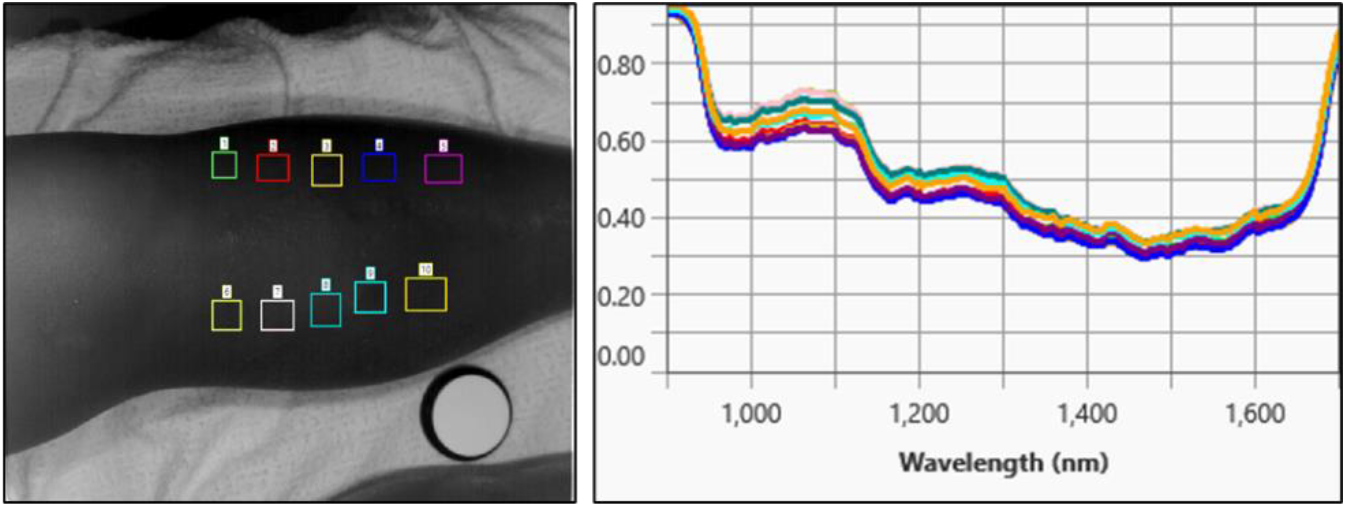
Overview of MCI data collection and processing.

For each subject, one hypercube was collected per shin. Every pixel in a hypercube contains a spectrum. In Figure 2C, an acquired shin image is overlayed with colored boxes indicating different regions of interest (ROIs) where spectral measurements are made, and the average reflectance spectra from each of the ROIs is shown. Spectra corresponding to soft tissue regions of the shin are extracted from the hypercubes and applied to multivariate statistical and classification methods, such as Partial Least-Squares Regression (PLSR).^21^ The PLSR model generates, from the spectra, regression vectors that are used to generate a SWIR-based prediction index (the CardioVerification Index, CVI) for any given spectrum (pixel). This is the measure of interstitial water beneath the patient’s skin. The CVI ranges from 0 to 1, to correspond to the MRI Dixon score of 0 to 1 for WF. Due to MRI analysis being restricted to only the medial side of the shin, for the CV analysis, ROIs were also defined on the medial side of the shin that corresponded to the same location as the MRI axial slices, and spectra from these regions were analyzed to determine their CVIs. Additional details regarding CV technology have been previously published. ^20^

**Figure 2C:**
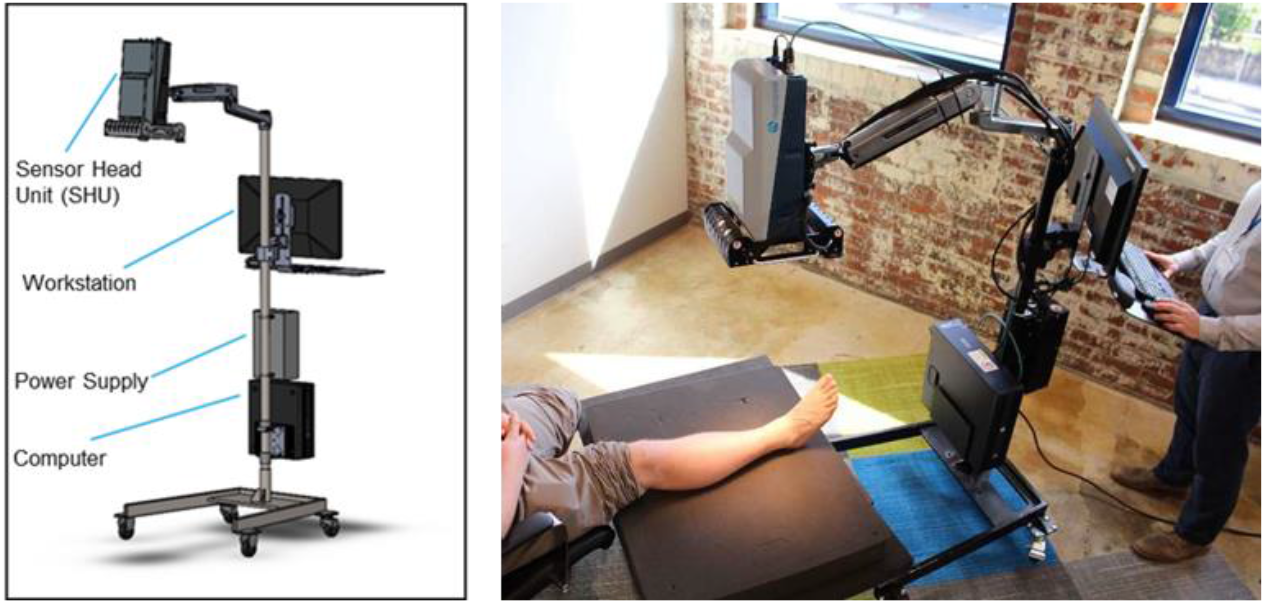
Left: Acquired shin image with colored ROIs where spectral measurements are made. Right: Average reflectance spectra from the ROIs (each colored trace matches the ROI of same color).

## Results

### Subject population

Forty-seven (47) subjects were enrolled, of whom 36 were ADHF inpatients and 11 were non-HF outpatients. All completed the required examinations and imaging. Subject demographics are shown in

**Table 1.** As a control group, the non-HF outpatients had significantly less edema than the ADHF inpatients, trended toward a higher male predominance, were less likely to be diabetic or smokers. Subject Demographics

| Characteristic | All Subjects<br>(N = 47) | ADHF Inpatients<br>(N=36) | Non-HF Outpatients<br>(N=11) | P-value<br>ADHF vs Non-HF |
| --- | --- | --- | --- | --- |
| Age (yrs, mean) | 59.6 | 66.1 | 36.8 | <0.0001 |
| Age (yrs, median) | 64 | 67 | 32.5 | <0.0001 |
| Age (yrs, range) | 23 – 96 | 38 – 96 | 23 – 60 | n/a |
| Sex (% male) | 70 | 61 | 91 | 0.0778 |
| Pitting edema = 0 | 22 | 11 | 11 | <0.0001 |
| Pitting edema = 1+ | 12 | 12 | 0 |  |
| Pitting edema = 2+ | 10 | 10 | 0 |  |
| Pitting edema = 3+ | 3 | 3 | 0 |  |
| Pitting edema = 4+ | 0 | 0 | 0 |  |
| BMI (avg) | Unknown | 31.9 | Unknown | n/a |
| % Diabetic | 36.9 | 48.5 | 0 | 0.0034 |
| % Smokers | 17.4 | 22.9 | 0 | 0.1691 |
| % Venous insufficiency | 6.5 | 5.7 | 9.1 | 1.0000 |
| % LV EF (mean) | Unknown | 45.7 | Unknown | n/a |
| % LV EF (range) | Unknown | 10 – 76 | Unknown | n/a |
| % HFpEF (EF > 40%) | Unknown | 58.8 | Unknown | n/a |

### Physical examination reliability

Twenty-nine (80%) of the ADHF inpatients underwent pitting edema grading by two independent clinicians. (None of the non-HF outpatients had pitting edema.) For all ADHF subjects except one, the two pitting edema grades were assigned within minutes of each other (6.9 ± 4.3 (μ ± σ) minutes). For the outlying subject, the two grades were assigned within two hours of each other.

Chi-squared analysis showed that there was a statistically significant difference between the clinicians’ assigned edema grades per subject (p=0.0002), and Cohen’s kappa showed that inter-rater agreement was fair-to-moderate (0.42).

### Water fraction measurement using the MRI Dixon sequence and CardioVere in the lower extremity

Because each subject had 4 independent MRI water fraction (WF) measurement sites (2 on the left shin and 2 on the right shin), 188 MRI WF measurements were taken. Applying the Dixon sequence to MRI images of the lower extremities posed no technical issues regarding image capture or display. Likewise, capturing SWIR images from the lower extremities posed no technical issues.

For 4 subjects, however, the reading radiologist determined that there was one or more region(s) of the tibia with insufficient subcutaneous fat to select an ROI for the Dixon sequence. By directing the CV operator to avoid those regions while performing SWIR imaging, opportunities for data capture were maximized. Notwithstanding this effort, twenty-one measurements (11.2%) were removed from the dataset because they were collected from mismatched MRI/SWIR imaging regions or shins affected by gross visual distortion (*i.e*., varicose veins, prior saphenous vein harvest) that prevented matching. This left 167 MRI WF/CVI WF collocated imaging pairs for analysis.

For these matched pairs, we found that both the MRI WF measurements and the CVI WF measurements were able to differentiate AHDF from non-HF subjects, although there was significant overlap between the two subject populations, for both groups (Figure 3).

**Figure 3:**
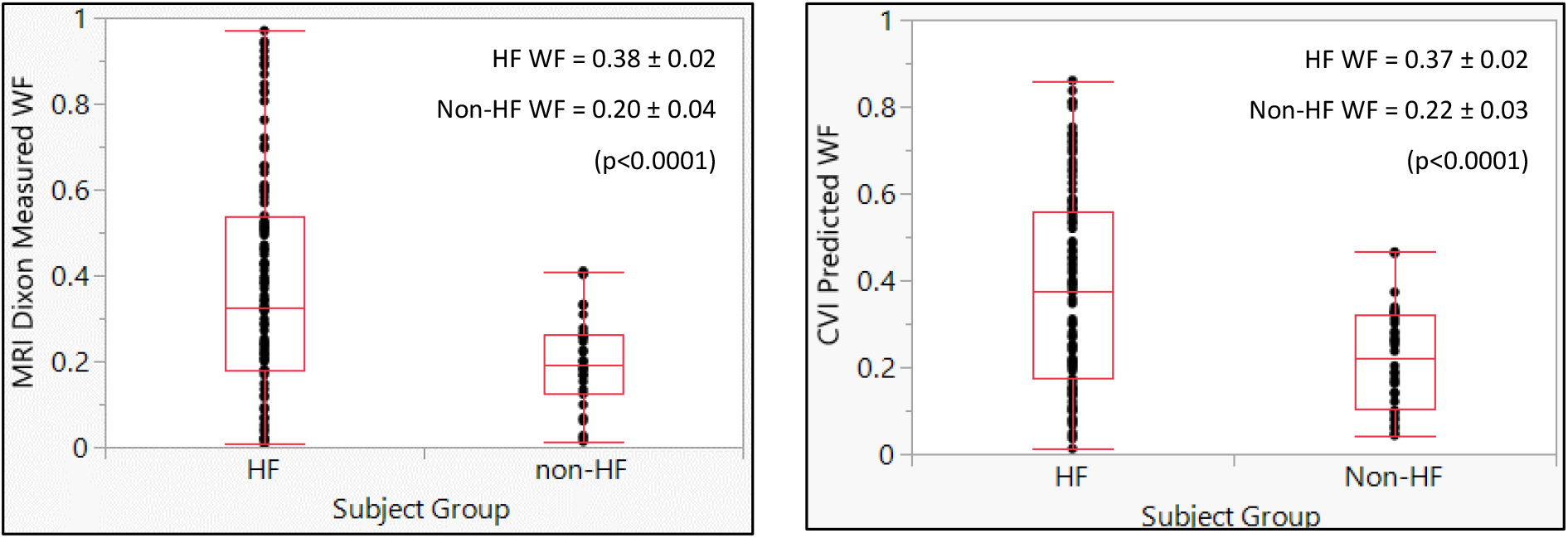
MRI WF (left) and CVI WF (right) Differentiate Subjects with ADHF from Subjects without HF

Some degree of overlap in WF measurements between the subject groups was expected, as was the observation that some ADHF subjects did not have appreciable TC at all. However, we were surprised to observe that some of the non-HF subjects demonstrated appreciable TC without any physical findings. This raised the possibility of using CVI to detect preclinical TC and potentially to screen for HF, which would identify TC in patients without signs or symptoms of the disease. We therefore present below our continued data analysis conducted along two lines: one analysis for all study subjects, and another for only those subjects with “Low TC”, that is, subclinical edema (pitting edema = 0) or very low edema (pitting edema = 1+).

### Comparing water fraction measurements between MRI and CardioVere

For each matched location on the shin, the PLSR model created to predict WF (the “CVI WF” model) output numerical values that differed somewhat from the measured MRI WF. The scatterplots visualizing the correlation between the MRI measured WF and the CVI predicted WF are shown in Figure 4, both for the entire subject population (left) and the low TC subpopulation (right). For both instances, there was excellent linear correlation between MRI WF and CVI WF (r^2^ = 0.743 and 0.674, respectively). Across all study subjects the MRI WF measurements ranged from 0.00 – 0.97; for the Low TC subjects the range was 0.01 – 0.72. The CVI WF predictions ranged from -0.09 – 0.88 for the entire subject population, and from 0.00 – 0.63 for the Low TC subpopulation.

**Figure 4:**
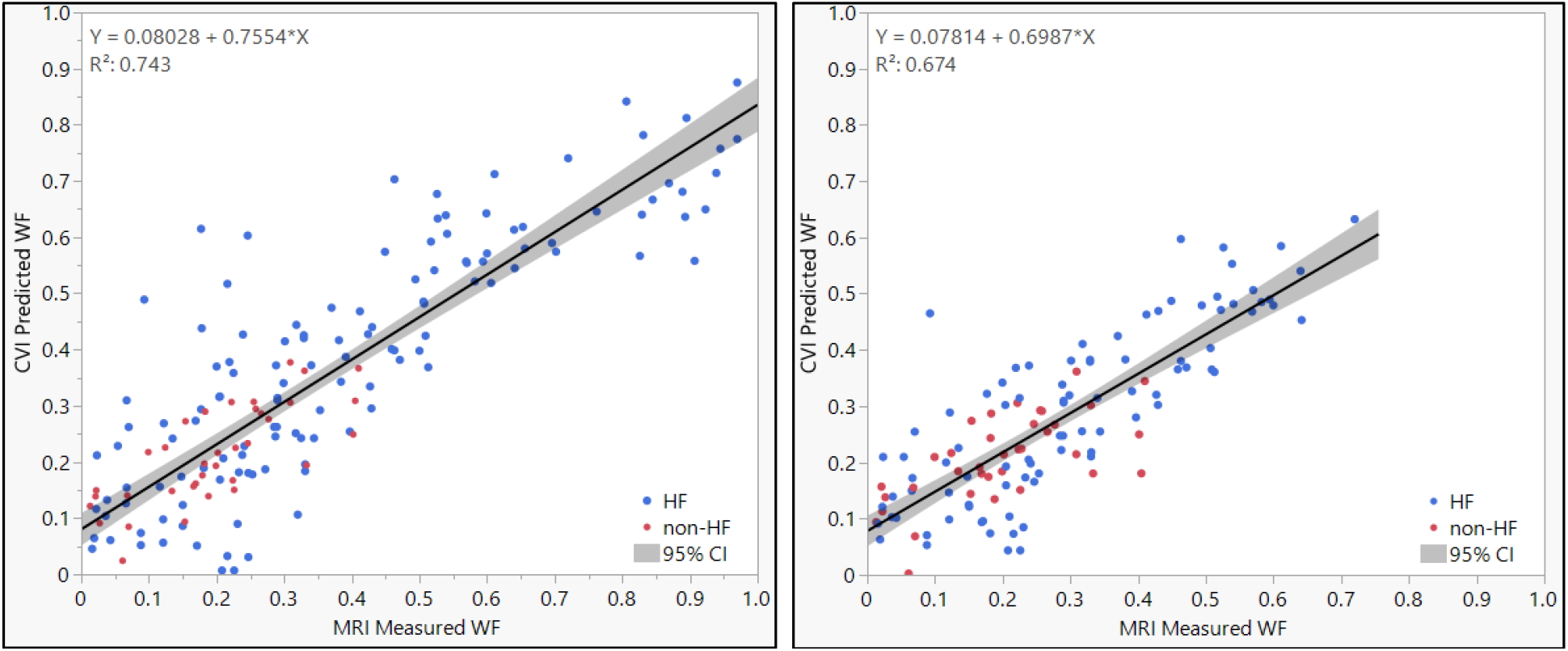
Scatterplots of Measured MRI WF and Predicted CVI WF for each matched imaging location on the shin. (Left) Total subject population. (Right) Low Tissue Congestion population only.

The overall performance evaluation regarding each CVI WF prediction model’s ability to differentiate HF subjects from non-HF subjects, both for the entire subject population and the Low TC subpopulation, is shown in Figure 5. The areas under the receiver operating characteristic curves (AUC) are 0.906 and 0.821 for the entire subject population and the Low TC subpopulation, respectively.

**Figure 5:**
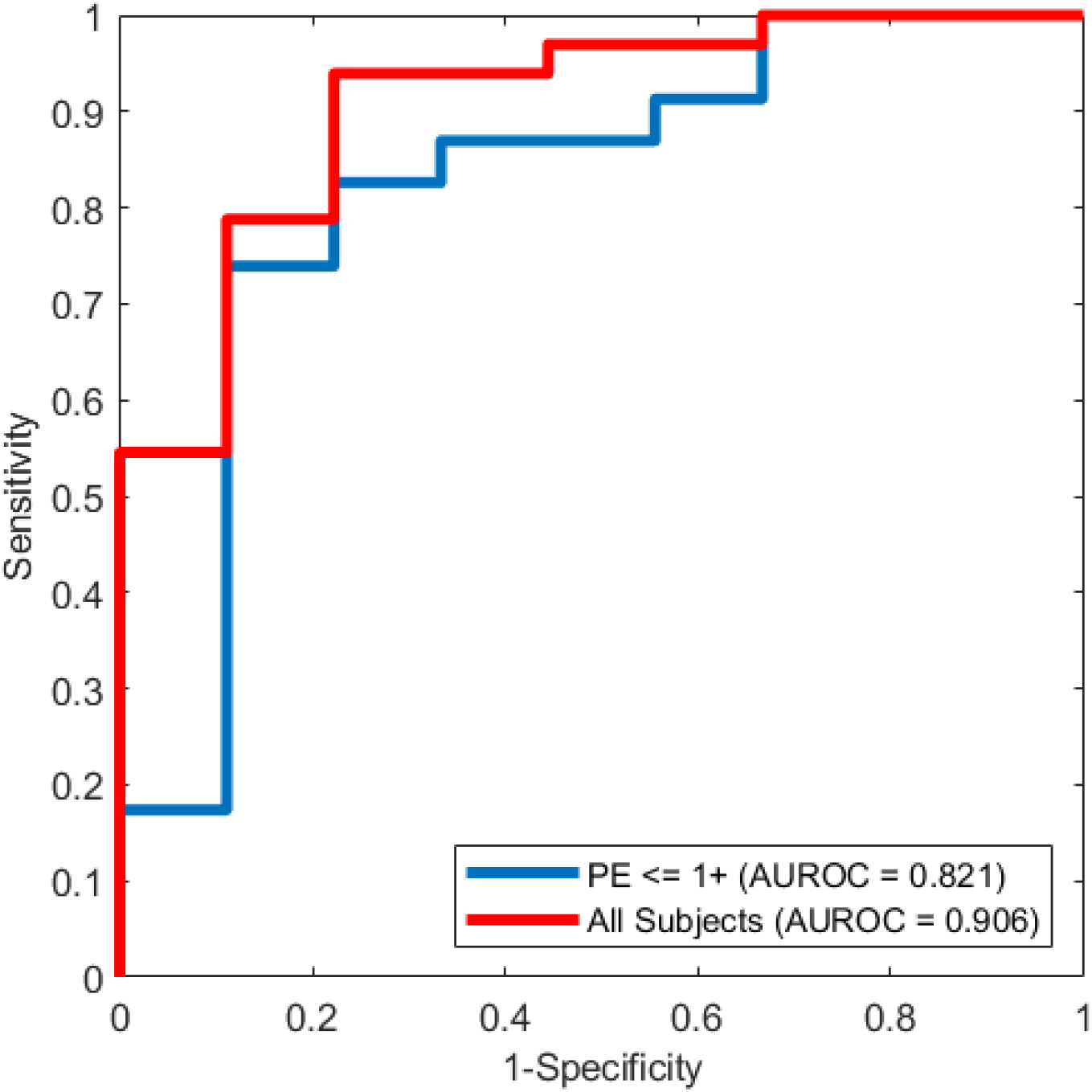
Receiver Operating Characteristic curves for the two CVI WF prediction models, showing ability to differentiate HF subjects from non-HF subjects. (Red) All study subjects; (Blue) Low Tissue Congestion subjects only AUROC = Area under the receiver operating characteristic curve

## Discussion

### Using the MRI Dixon sequence method in the lower extremities

The Dixon sequence for MRI studies, developed more than thirty years ago, has been used widely to measure fat content in solid organs. To our knowledge, it has not been used to measure water content in the periphery. We have demonstrated in this study that the Dixon sequence method can be used successfully to quantitatively measure interstitial water in the lower extremity.

Although not presented in this paper, during this study we also captured MRI and SWIR MCI images from subjects’ proximal lower extremities (thighs) and captured SWIR MCI images from forearms. Analysis is currently in process with the intent of showing that these measurements also accurately reflect interstitial water content. These results will be included in a forthcoming publication. If successful, the proven ability to accurately measure TC in patients’ thighs and/or forearms will open the door to overcoming the ergonomic challenges many HF patients face when taking images from certain parts of their bodies, such as the shins.

### Using pitting edema to guide HF therapy

In this study we confirmed the work of others that even when two highly trained and experienced clinicians assess TC by grading pitting edema, and that even when those assessments are made only minutes apart, there is still a high degree of variability and inter-rater disagreement. Now that MRI and CVI can be used to measure TC objectively and reproducibly, we argue that pitting edema should no longer be used in clinical practice, neither to guide the care of HF patients in the clinic nor as a ground truth for the development of HF detection, management algorithms or care protocols.

### The role of tissue congestion in HF screening

Tissue congestion is the common pathophysiologic pathway for all patients with HF, regardless of etiology. This suggests that screening for HF should begin with detecting TC, if TC is in fact present prior to overt HF symptomatology. However, no screening modality has yet been directed at detecting and quantifying TC in the asymptomatic patient. The two most widely evaluated modalities to date, echocardiography and natriuretic peptide levels, both suffer from significant detractions:

- Echocardiography images cardiac anatomy and function. It can detect diminished pump function and/or valvular disease but not TC. It is also expensive, not widely available, requires complex analysis to detect patients with HF and a preserved ejection fraction, and is highly dependent upon operator expertise.^22–24^
- Elevated natriuretic peptide levels have been shown to increase the post-test probability of HF in certain patient populations, but the results are confounded by very high false positive rates and a wide range of rule-in and rule-out values.^25^ Testing is also invasive, costly, and not real-time.

Our discovery that WF measurements were elevated not only in ADHF subjects with low edema on exam, but also in non-HF subjects with no edema, strongly suggests that “preclinical” TC is present before detection on physical exam is possible. (Indeed, it may be true that some of the “non-HF” subjects were undiagnosed stage B HF patients.) This ability of CVI to detect preclinical TC means that the device fits perfectly the definition of a HF screening test, and once the technology advances to a handheld, portable, inexpensive device, it could become a standard of care to screen at-risk HF patients with CVI, bringing undiscovered stage B patients into early treatment.

This proposition is strongly supported by the ROC curves shown in Figure 5, where CVI demonstrates excellent performance at differentiating HF from non-HF subjects, exactly the use case that fits in-office screening for asymptomatic, at-risk HF patients.

### The role of tissue congestion in monitoring HF

Readmission rates after hospitalization for acute HF decompensation remain unacceptably high, despite continued advances in drug development and guideline-directed medical therapy protocols.^26^ Prime targets for improvement are such factors as patient non-compliance and poor monitoring in the outpatient setting.^27–29^ By making CVI available to patients and their caregivers at home on an inexpensive, easy-to-use platform, the barrier to routine monitoring and compliance will diminish. By connecting CVI to a digital platform, early discovery of increasing TC will enable patients and their clinicians to intervene early and halt decompensation. Future work is needed to understand the utility of CVI in longitudinal care settings.

### The role of tissue congestion in managing ADHF

Another cause of high readmission rates may be too-early or too-late hospital discharge after an episode of acute HF decompensation. Clinicians presently do not have a quantitative, objective measure or constellation of measurements to which they can refer – pulmonary congestion resolution marked by absent crackles, peripheral congestion resolution marked by absent pitting edema or reduced body weight are all highly subjective and often lagging indicators of interstitial fluid status.

We have found previously in healthy subjects that SWIR MCI can track changes in TC over time (unpublished results), and in this study we have shown that the CVI WF is a valid method of TC in the hospital. The next step therefore is to determine whether using CVI WF over time in hospitalized ADHF patients can help clinicians determine when a particular patient has been diuresed sufficiently for hospital discharge with minimal risk of early readmission.

### Study limitations

We found that neither the MRI/CVI relationship for all subjects nor the MRI/CVI relationship for low-congestion subjects demonstrated a perfect linear correlation or a slope of exactly 1.0, although the prediction accuracy of both CVI models to differentiate between HF and non-HF subjects was high. This suggests that there is still unexplained variability in the subject population regarding measuring TC, and that further analysis and modeling is required. One area of exploration would be the creation of separate models for low-congestion versus high-congestion situations, and this awaits further research.

Another limitation is that we have not yet defined what is a “normal” CVI. This will require studying subjects who have normal cardiovascular physiology and fluid status compared to subjects who are congested and fluid overloaded. From these data we will be able to calibrate CVI and help inform clinicians whether a patient they are screening likely has HF or not.

Finally, we are limited in our interpretation because our subject population likely does not represent the overall population of patients who are at-risk for HF or who have HF. For example, our subject group was overwhelmingly Caucasian, whereas a disproportionate number of African American and Hispanic patients suffer from HF.

### Conclusions

In this validation study, we utilized the MRI Dixon sequence in a novel manner and found that it can quantify tissue congestion in the lower extremities of patients with HF. We also noted that in patients hospitalized for HF, CV assessment of subcutaneous water fraction was feasible and correlated with measurement by MRI. According to both MRI and CVI, there was a broader range of congestion present in the participants than was appreciated clinically. We also concluded that CV could detect and quantify TC in the lower extremities in both clinical and subclinical conditions and accurately differentiate between patients with and without HF.

The study also showed that pitting edema is not a clinically useful measure of TC. Our study demonstrated that CV has the potential to help clinicians manage heart failure patients in the hospital as well as screen asymptomatic patients in stage A and stage B who may be at high risk for HF or may have HF in its earliest stages where clinical signs are not evident. As our next step, we need to validate these findings in a prospective, randomized, double-blinded trial across the entire spectrum of treatment settings such as outpatient clinics, hospital admissions and monitoring clinics. We also aim to explore the accuracy of CV in detecting TC across different patient body habitus and skin shades.

## Data Availability

All data produced in the present study are the property of ChemImage.

## Acknowledgments

The authors acknowledge Siemens USA (Washington, DC) for their gracious and generous provision of their LiverLab software to calculate MRI water fractions. The authors also acknowledge ChemImage Corporation (Pittsburgh, PA) for funding the study and providing the CardioVere prototype.

